# Identifying novel gene dysregulation associated with opioid overdose death: A meta-analysis of differential gene expression in human prefrontal cortex

**DOI:** 10.1101/2024.01.12.24301153

**Authors:** Javan K. Carter, Bryan C. Quach, Caryn Willis, Melyssa S. Minto, PGC-SUD Epigenetics Working Group, Dana B. Hancock, Janitza Montalvo-Ortiz, Olivia Corradin, Ryan W. Logan, Consuelo Walss-Bass, Brion S. Maher, Eric Otto Johnson

## Abstract

Only recently have human postmortem brain studies of differential gene expression (DGE) associated with opioid overdose death (OOD) been published; sample sizes from these studies have been modest (N = 40-153). To increase statistical power to identify OOD-associated genes, we leveraged human prefrontal cortex RNAseq data from four independent OOD studies and conducted a transcriptome-wide DGE meta-analysis (N = 285). Using a unified gene expression data processing and analysis framework across studies, we meta-analyzed 20LJ098 genes and found 335 significant differentially expressed genes (DEGs) by OOD status (false discovery rate < 0.05). Of these, 66 DEGs were among the list of 303 genes reported as OOD-associated in prior prefrontal cortex molecular studies, including genes/gene families (e.g., *OPRK1, NPAS4*, *DUSP, EGR*). The remaining 269 DEGs were not previously reported (e.g., *NR4A2, SYT1, HCRTR2, BDNF*). There was little evidence of genetic drivers for the observed differences in gene expression between opioid addiction cases and controls. Enrichment analyses for the DEGs across molecular pathway and biological process databases highlight an interconnected set of genes and pathways from orexin and tyrosine kinase receptors through MEK/ERK/MAPK signaling to affect neuronal plasticity.

## Introduction

The opioid epidemic continues to be a tremendous burden on our society and communities around the world. In the United States, 5.6 million people ages 12 and older misused opioids during 2021 (1,2). In the same year, the United States saw the highest 12-month count of opioid overdose deaths (OOD) recorded, >80LJ000, a 40% increase since 2019 (1). Although our understanding of the neurobiology of opioid addiction remains limited, increased sample sizes from recent meta-analyses of genome-wide association studies (GWAS) have begun to identify genetic loci robustly associated with opioid addiction, including loci in or near *OPRM1*, *FURIN*, and *SCAI/PPP6C/RABEPK* (3–5). However, such robust findings for critical features of gene regulation in the human brain, and gene expression in particular, have yet to emerge.

Only recently have human postmortem brain studies of differential gene expression (DGE) associated with OOD been published: Corradin et al. 2022 (6); Mendez et al. 2021 (7); Seney et al. 2021 (8); and Sosnowski et al. 2022 (9). All four of these studies used human postmortem dorsolateral prefrontal cortex (DLPFC) brain tissue from donors identified as dying from OOD through toxicology assays administered by forensic scientists and phenotypic evidence of opioid addiction. Each of these independent studies had modest sample sizes (N = 40-153) and compared bulk RNA-seq data from individuals who died from OOD to individuals who died from non–drug use causes. The DLPFC region of the brain involves the preoccupation/ anticipation component of the addiction cycle, which affects craving, impulsivity, and executive function (10). The DLPFC has also been linked to DGE levels studies of *OPRM1* (11) and associated with anxiety and impulsive neurological disorders (ADHD, schizophrenia, bipolar disorder, etc.). Addictive and reward-response phenotypes are also traits reported to be linked to the DLPFC from human and animal model studies, which can lead to drug-seeking behavior. Each DLPFC DGE study reported OOD-associated genes with plausible biological links to addiction. However, with their sample sizes, statistical power to robustly identify OOD-associated DGE is limited. Here, we uniformly processed the bulk RNA-seq data across the four OOD studies and performed a DGE meta-analysis including 283 samples (case N = 170, control N = 113), making this the largest transcriptome-wide analysis of OOD to date. Our findings include novel differentially expressed genes (DEGs) in addition to confirming a subset of previously reported OOD-associated genes. We link our meta-analysis DEGs to different biological processes and pathways, notably the orexin receptor signaling system and signaling by Receptor Tyrosine Kinases. Furthermore, we investigated genetic regulation of OOD-associated DEGs and assessed shared genetics between these genes and 47 GWAS traits through partitioned heritability and colocalization analyses (12–14).

## Methods

### Contributing Study Cohorts

Characteristics of the four cohorts contributing to this meta-analysis are provided in Table 1. Each of the contributing studies defined cases as decedents whose death was attributed to opioid overdose based on toxicological analyses by corners’ offices and phenotypic evidence of a history of opioid misuse or opioid addiction. Corradin study: Used de-identified human cadavers where cause of death was defined from “forensic pathologist following medico-legal investigation evaluating the circumstances of death including medical records, police reports, autopsy findings and forensic toxicology analysis” (6). Mendez study: Postmortem brains were collected from the University of Texas Health Science Center at Houston Brain Collection during routine autopsies, with approval from the Institutional Review Board. Detailed psychological autopsies were conducted, including information on psychiatric phenotypes, age of onset of drug use, types of drugs used, and co-morbidities. All but one sample was positive for opioids on toxicology at time of death (7). Seney study: Samples were obtained during routine autopsies by the Office of the Allegheny County Medical Examiner after obtaining consent from next-of-kin. Autopsy and toxicology analyses were conducted, and subjects with OUD were matched with unaffected comparison subjects for sex and age. Subjects with OUD had a diagnosis duration of 5-18 years (8). Sosnowski samples: Postmortem brain samples were donated to the Lieber Institute for Brain Development from the Offices of the Chief Medical Examiner, with detailed neuropathological examinations and retrospective clinical diagnostic reviews for psychiatric and medical historiesAll brain donors had forensic toxicological analysis, “which typically covered ethanol and volatiles, opiates, cocaine/metabolites, amphetamines, and benzodiazepines” (9).

**Table 1.**
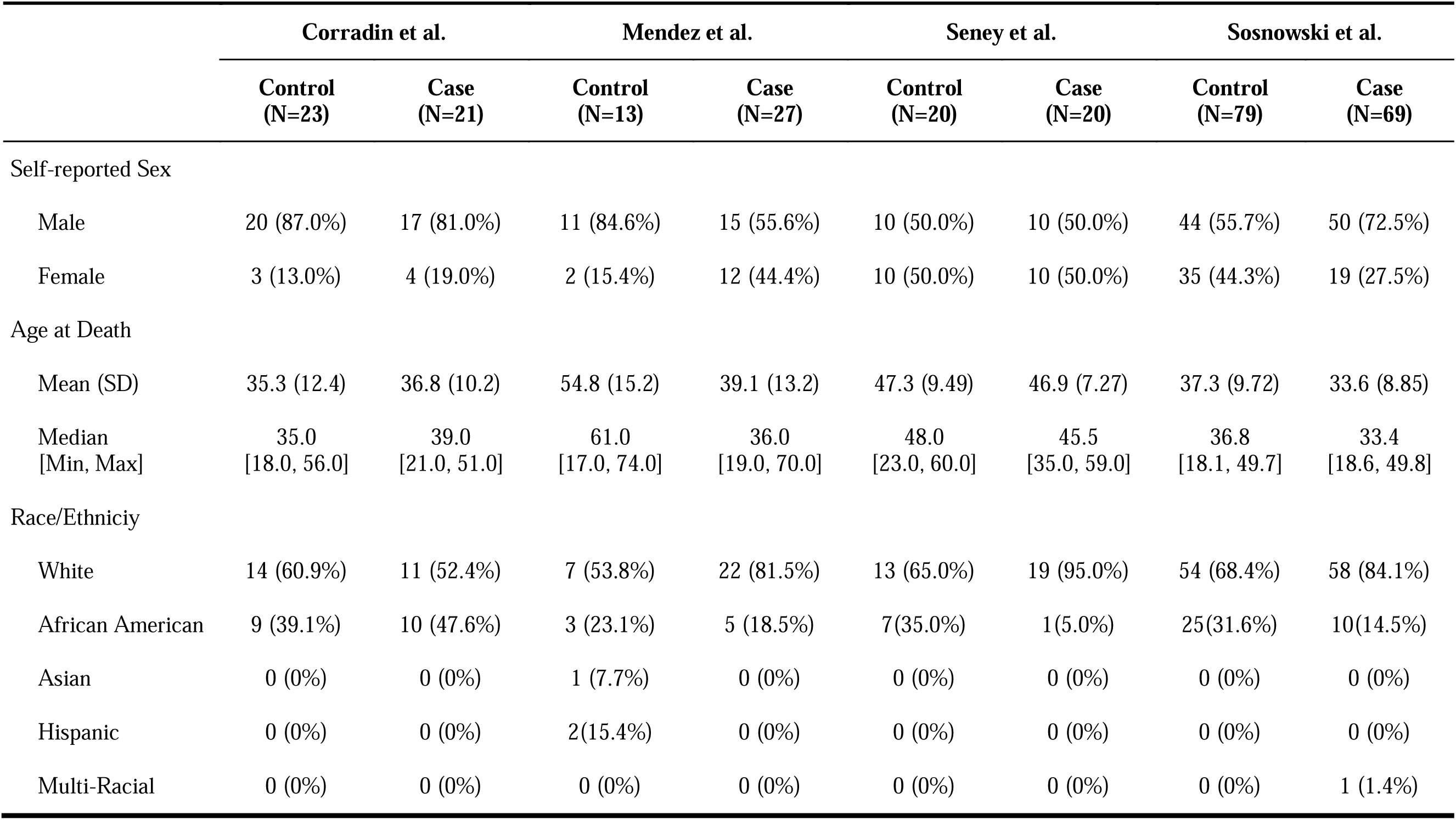
Samples from all four studies used in the meta-analysis.

### RNA Sequencing and Data Processing

Paired-end RNA-seq FASTQ files were obtained from four publicly available datasets (Sequence Read Archive study ID SRP324812, Sosnowski et al. (9), Gene Expression Omnibus accession IDs GSE174409, Seney et al. (8) and GSE182321, Mendez et al. (7), and dbGaP study phs002724.v1.p1, Corradin et al. (6)) and processed through a unified workflow. Briefly, adapters were trimmed and reads filtered using *Trimmomatic* v0.39 (15). Reads were then pseudo-mapped using *Salmon* v1.1.0 (16) in selective alignment mode (17) using the GENCODE v30 comprehensive gene annotation as the transcriptome index and the full GRCh38 primary genome assembly as a selective alignment decoy sequence. Salmon transcript quantifications (mapping percentage > 30%) were aggregated to the gene level using *tximport* v1.12.3 (18). Quality metrics for raw and post-*Trimmomatic* reads (retained reads percentage > 60%) were generated using *FASTQC* v0.11.8 (19), and reads were aligned to the GRCh38 genome using *HISAT2* v2.1.0 (20) to generate additional quality metrics. All quality metrics and read mapping statistics were aggregated using *MultiQC* v1.7 (21). Samples were then filtered based on several quality control criteria (e.g., effective sequencing depth > 10M, read GC content between 35% and 65%, transcriptome mapping perfectage > 50%, RNA integrity number (RIN) score >=5, mitogenome mapping percentage < 50%, and Ribosomal RNA mapping percentage < 1%.

### Differential Gene Expression Analysis

For each dataset, a regression model was fit using *limma* with voom-transformed count data (22). Prior to model fitting, lowly expressed genes were removed (< 10 gene counts in the approximate proportion of samples that comprise the smaller OOD status group). To account for dataset-specific characteristics, covariates include in the regression model for each dataset reflected those from their respective published studies. All models included OOD status, age, sex, post-mortem interval (PMI), and RIN as covariates. Additionally, the Corradin model included race, sequencing batch, and 12 surrogate variables estimated by SVA (23). The Mendez model included cerebellar pH. The Seney model included race and brain tissue pH. The Sosnowski model included race, cocaine/amphetamine toxicology report status, ribosomal RNA mapping rate, gene assignment rate, mitochondrial RNA mapping rate, concordant read pair mapping rate, overall mapping rate, External RNA Control Consortium spike-in error rate, and 10 quality control surrogate variables.

### Meta-analysis

To combine evidence of differential expression across datasets, we use the samples sizes and differential gene expression analysis p-values from our independent analyses of each study to conduct a weighted Fisher’s meta-analysis implemented by the R package *metapro* (24). Any genes that were only tested in one study (because they were lowly expressed in the others) were removed, and a Benjamini-Hochberg FDR threshold of < 0.05 was applied to declare a gene as significantly differentially expressed.

### Gene Set Overrepresentation Analysis

The gene set overrepresentation analysis was conducted using the ToppFunn tool from ToppGene Suite, Transcriptome, ontology, phenotype, proteome, and pharmarcome annotations-based gene list functional enrichment analysis (Toppfun) (25) to identify GO terms, biological pathways, and disease-annotated gene sets enriched for meta-analysis DEG. GO terms included gene sets from the MF, biological process, and cellular components categories (26,27). Biological pathways were from the MsigDB C2 BIOCARTA collection (v7.5.1) (28–30). Disease-annotated gene sets were sourced from DisGeNET (BeFree &Curated) (31) (Supplementary Table 1). The R package *simplifyEnrichment* (32) was used to do semantic similarity-based hierarchical clustering of GO biological process terms using the Wang et al. (33) distance metric (Figure 2.A). We set the seed in Rstudio for “888” for constant results (i.e., set.seed(888)). Binary cut was used to determine GO term clusters. Nine other clustering methods were compared against the binary cut method (Supplementary Figure 1), but were outperformed in similarity scores and cluster quantity by binary cut.

### Partitioned Heritability Analysis

In our study, we utilized sLDSC (34) to assess the heritability of 47 GWAS traits captured by the genomic regions spanning the OOD-associated differentially expressed genes (DEGs). Of these traits, 41 have been previously examined in relation to OUD (5) and 6 in relation to sleep (i.e., insomnia and differential sleep duration) (35,36) (Supplementary Table 2) For each of the 335 meta-analysis DEGs, we extracted the genomic region encompassing the gene body and 100 kilobases upstream and downstream of the gene ends. These regions, along with GWAS summary statistics for the 47 traits, were provided as input for sLDSC. We utilized the GENCODE v30 (17) comprehensive gene annotation GTF file for GRCh37 to obtain gene start and end coordinates and converted the GWAS summary statistics for each trait to the build GRCh37 format. An LD reference panel derived from the 1000 Genomes Phase 3 EUR superpopulation was used for the LD scores. The munge_sumstats.py script from the LDSC GitHub repository was used to ensure consistency in the formatting of the GWAS summary statistics.

#### Differential Cell-type Proportion Testing

Cell types were inferred using the BISQUE deconvolution tool (37) using DLPFC single-nuclei RNA-seq (38) as the reference panel. The proportion of the nine cell types present in the reference (astrocytes, GABAergic neurons, excitatory neurons, macrophages, microglia, mural cells, oligodendrocytes, oligodendrocyte precursors, and T-cells) were estimated in the bulk RNA-seq data from the four studies. To determine whether cell type proportions differed by OUD status for each study, cell-type proportions were arcsin transformed, and a linear regression model was fit for each cell type with the transformed proportions as the outcome variable. The explanatory variables for all models included age, sex, RIN, PMI, and OOD status. For each study, additional covariates were included (Corradin: RNA-seq batch and Race; Mendez: cerebellar pH; Sosnowski: cocaine amphetamine tox, “rRNA_rate,” “concordMapRate,” “overallMapRate,” “ERCCsumLogErr,” race). Two-sided t-tests were conducted within each dataset to assess study-specific associations between OOD and cell type proportion. Finally, a Fisher’s meta p-value was calculated by using the p-values from the sumlog function from metap v1.8 R package (39). Cell-type proportions were considered as significantly different by OOD status for Bonferroni-adjusted meta-analysis p-value < 0.05 (Supplementary Table 3).

#### Study Reported Gene List

DEGs from the prior studies were identified for our purposes based on the thresholds used and significance reported by each respective study and concatenated (N = 303): Corradin et al., 2022; Bonferroni corrected p-value < 0.05 (N = 10); Mendez et al., 2021; FDR p-value < 0.05 and |FC| >1.5” (N = 29); Seney et al., 2021; FDR p-value < 0.01 and log_2_ FC > ± 0.26 (i.e., FC ± 1.2 or 20% expression change), focused on the top 250 genes of 567 (N = 250); Sosnowski et al., 2022; FDR corrected p-value < 0.10 and log_2_ FC < -1.5, with the addition of three extra genes (N = 4).

### Expression Quantitative Trait Loci Look-up

We copied and transferred the summary statistics from the GTEx portal single-tissue cis-QTL dataset (67,68) “Brain_Frontal_Cortex_BA9.v8.signif_variant_gene_pairs.txt” and “Brain_Forntal_Cortex_BA9.v7.signif_variant_gene_pairs.txt” for the colocalization analysis to match the GRCh37 reference genome used in the opioid addiction GWAS. Next, we used a custom script that matched the ensembl ID from the meta-analysis 335 gene list and matched them in the eQTL summary statistics text file. The matched eQTL’s p-values threshold was < 0.05.

### Colocalization of eQTL Egenes and Observed Traits Associated to OOD

Colocalization was tested using the coloc v3.2.0 R package (42) with Brain Frontal Cortex (BA9) eQTL summary statistics from the GTEx Portal Version 7 for variants within 1 Mb of the meta-analysis DEGs and summary statistics for 35 GWAS phenotypes (Supplementary Table 4, a subset of phenotypes used for the partitioned heritability analysis. All summary statistics used were in Genome Build GRCh37/hg19. Using the coloc.abf() function, the eQTL data were specified as a quantitative trait while the GWAS data were specified as case-control for binary traits or quantitative otherwise. Only variants in both the eQTL summary statistics and the GWAS statistics were used (Supplementary Table 5). Colocalization hypothesis 4 posterior probability > 0.7 was considered as a significant colocalization.

## Results

### Differential Gene Expression Meta-Analysis

To increase statistical power and further our understanding of OOD, we leveraged the combined sample sizes of the four studies in a meta-analysis (N = 283, cases =170, controls = 113). We used the gene expression data generated from our uniform RNA-seq data processing workflow and fit regression models consistent with those used by the original studies (see Methods). Next, we combined resulting DGE summary statistics (yielding N = 20LJ098 genes) and combined p-values using the “metapro” wfisher function. Most of the tested genes were present in all four studies with the remaining tested genes occurring in two or three of the four studies (N = 20 098). Genes present in only one study were excluded from the meta-analysis (N=1 781; see Supplementary Figure 2). The results of the wfisher combined meta-analysis identified 335 significantly DEG (false discovery rate [FDR] < 0.05) as seen in Figure 1.A and 1.B. Of the 335 DEGs gene set, 85.2% were protein coding and 10.6% were long noncoding RNA genes (Figure 1.C). Comparing the list of 303 DEGs reported in the prior DGE studies with the 335 DEGs from this meta-analysis, we observe that 66 genes from the prior studies (6,8,9,43) were retained, including *DUSP2, DUSP4, DUSP6, EGR1, EGR4, ARC*, and *NPAS4* (Supplementary Table 6 & 7) (Figure 1.D).

**Figure 1.**
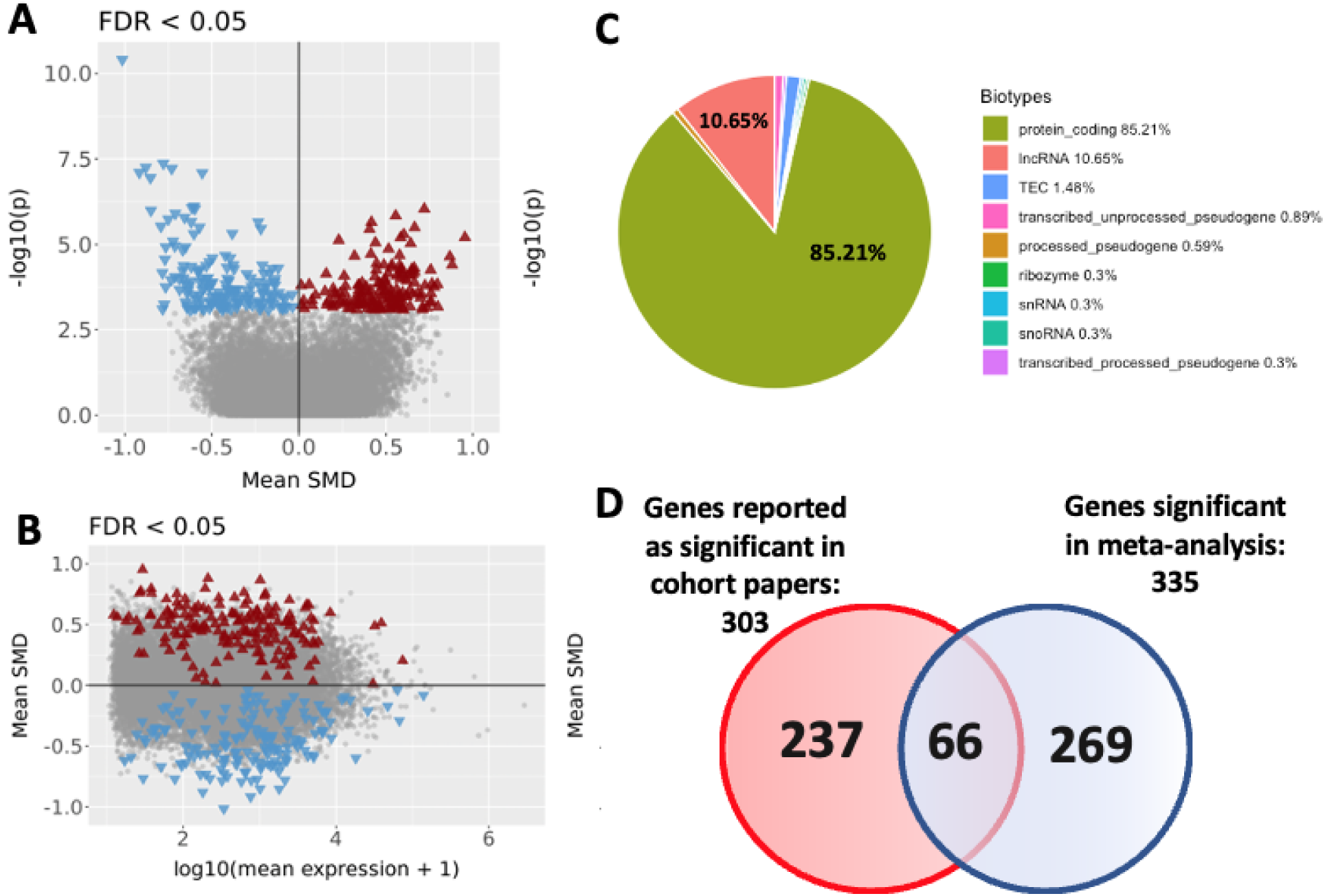
**A.** Volcano plot (FDR-BH corrected p-value < 0.05) for genes tested in the differential gene expression meta-analysis. Differential expression magnitude is represented here using the standardized mean differences (SMD) across studies. A total of 169 up-regulated (red triangle) and 166 down-regulated (blue triangle) significant genes; **Figure 1B.** MA plot (FDR-BH corrected p-value < 0.05) using the SMD; **Figure 1C.** Biotypes of the 335 significant genes from meta-analysis partitioned into percentages of the various different types of genetic biomarker types; **Figure 1D.** *Venn diagram representing how many genes were reported genes (significantly expressed genes documented in the four studies described above), meta-analysis genes (significant differentiated expressed genes Bonferroni corrected), and the 66 retained genes (number of genes that are present in both candidate gene list and meta-analysis gene list)*.

### Cell Type Deconvolution

Previous studies have indicated that opioid use can affect cell type proportion within the brain (8,44). To investigate cell type proportions in this meta-analysis, we used a single cell Tran et al. (38) reference dataset of the DLPFC to deconvolve the cell type proportions for each sample across the four independent datasets for nine cell types (astrocytes, GABAergic neurons, excitatory neurons, macrophages, microglia, mural cells, oligodendrocytes, OPCs, and T-cells) (Supplementary Table 3). Significant differences in microglia cell type proportions were observed within the Sosnowski dataset (Bonferroni p-value = 0. 0.002) and in the meta-analysis of cell type differences across all four studies (Bonferroni p-value = 0.039) (see Supplementary Figure 3). Although microglia proportions are significantly different between OOD case and control, our analysis shows varying direction of effect across cohorts and that the statistical significance in the meta-analysis is primarily driven by the Sosnowski et al. (9) results. Although limited by the reference panel for predicting cell type proportions in DLPFC, these results suggest that cell type proportions played a minimal role in the observed DGE.

### DEG Characterization Through Enrichment Analyses

#### Gene Ontology Terms

Gene Ontology (GO) enrichment analysis was performed on the significant meta-analysis genes to better understand the roles these genes may play in OOD. The 335 genes were tested for enrichment GO molecular function (MF), cellular component (CC), and biological processes (BPs), resulting in 78 significant terms (FDR-BH p-value < 0.05). Of the 78, three were GO MF terms relating to signal transduction and transcription factor activity including “MAP Kinase phosphatase activity” (p-value = 0.0412), “nuclear glucocorticoid receptor binding” (p-value = 0.0412), and “DNA-binding transcription factor activity, RNA polymerase 2 specific” (p-value = 0.0412). Ten of the enriched GO terms were GO CC terms and 65 were GO BP terms that reflected functions in synaptic signaling, chromatin regulation, and synaptic development, all of which are important in synaptic plasticity (45,46). Semantic clustering was performed on the enriched CC and BP terms separately. Within the GO CC terms, two clusters were identified: the largest being “vesicles dense cores,” which include synaptic vesicles followed by “acetyltransferase histone complex.” Of the GO BP terms, eight clusters were identified, including “morphogenesis,” “transcription regulation RNA,” “transsynaptic signaling synaptic,” “memory,” and “response hormone” with the largest z-score (Figure 2.A). The largest cluster of GO BP terms enriched were terms relating to morphogenesis, projection, and neuronal development.

**Figure 2.**
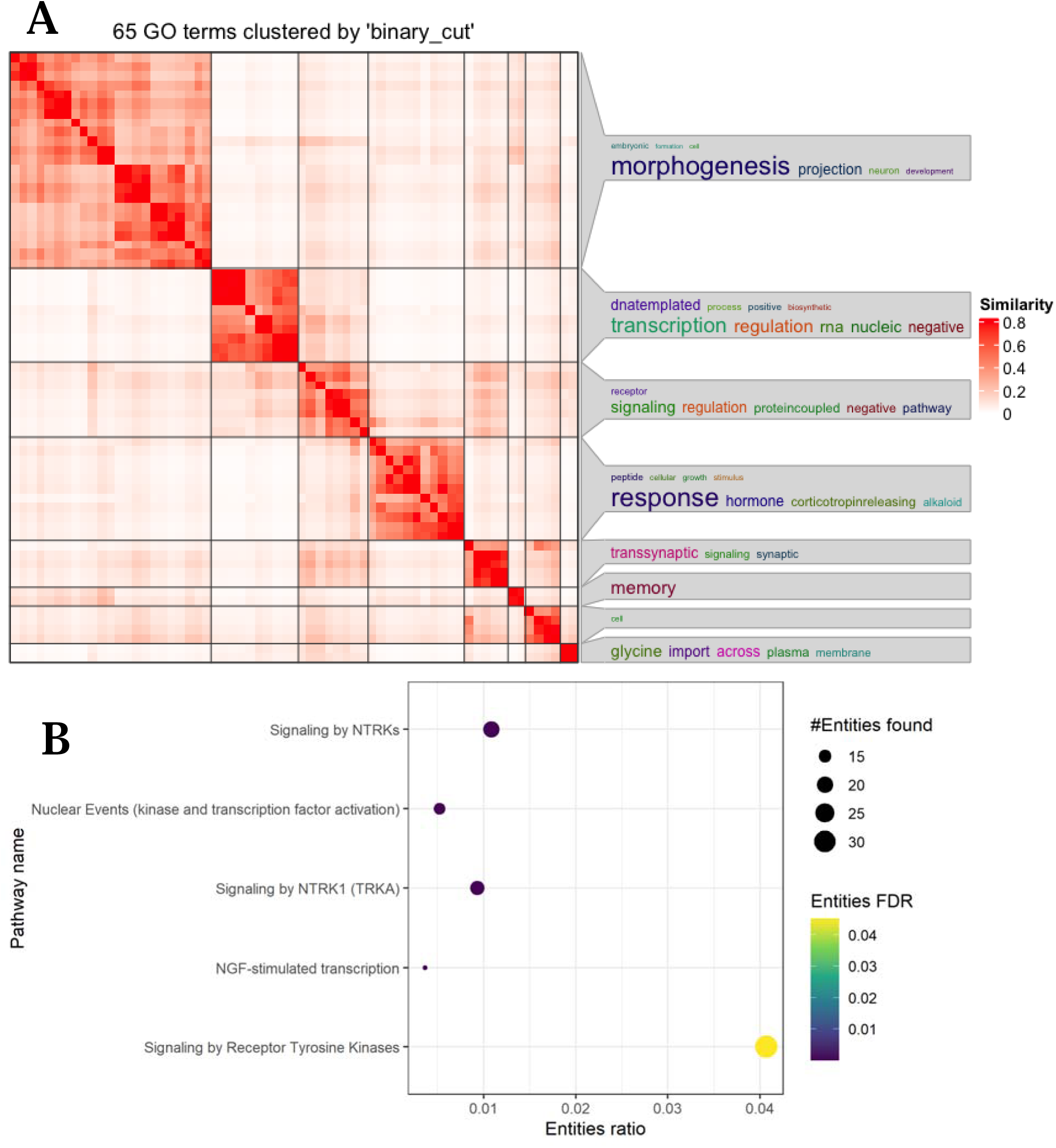
**A.** Heatmap of FDR significant p-values < 0.05 toppgene biological process GO terms N = 65, clustering of semantic similarity, the larger the characteristic, the greater the correlation. **Figure 2B.** Dot plot of Reactome databases results from differentially expressed gene list. Entities (nucleic acids, proteins, complexes, vaccines, anti-cancer therapeutics, and small molecules) participating in reactions form a network of biological interactions and are grouped into pathways.

#### Reactome Pathways

To expand biological characterization of the 335 DEGs, with a focus on relationships among signaling and metabolic molecules, we conducted a Reactome pathway enrichment analysis (47). Of the 335 submitted genes, 182 were present in the Reactome database. Five pathways were significantly enriched for our DEGs (FDR < 0.05; Figure 2.B). The primary pathway was Signaling by Receptor Tyrosine Kinase (RTK): 32 DEGs among the 625 in this pathway, entity ratio 0.04. The remaining four pathways are all nested under the primary pathway, with the enrichments driven by subsets of the 32 DEGs linked with the Signaling by RTK pathway, Signaling by NTRKs, Signaling by NTRK1, Nuclear Events (kinase and transcription factor activation, and Neuronal Growth Factor stimulated transcription. Signaling by RTK is itself a subfamily of the Signal Transduction pathway, constituting a class of cell surface proteins enabling ligand binding for stimulating intracellular cascades. Among other functions, these TRK pathways mediate synaptic plasticity that may be affected by opioid addiction or OOD (Supplementary Table 8 & 9).

When comparing the complete meta-analysis gene list (N = 335) to the retained gene list (N = 66) via GO similarity clustering, disease association (presented below), and pathway association, an increase in gene count within said associations was observed (i.e., biological process GO; Figure 3.A. This result indicates a strengthening of these GO term enrichments with the increase in statistical power under the meta-analysis (Supplementary Figure 1) (Figure 3.A).

**Figure 3.**
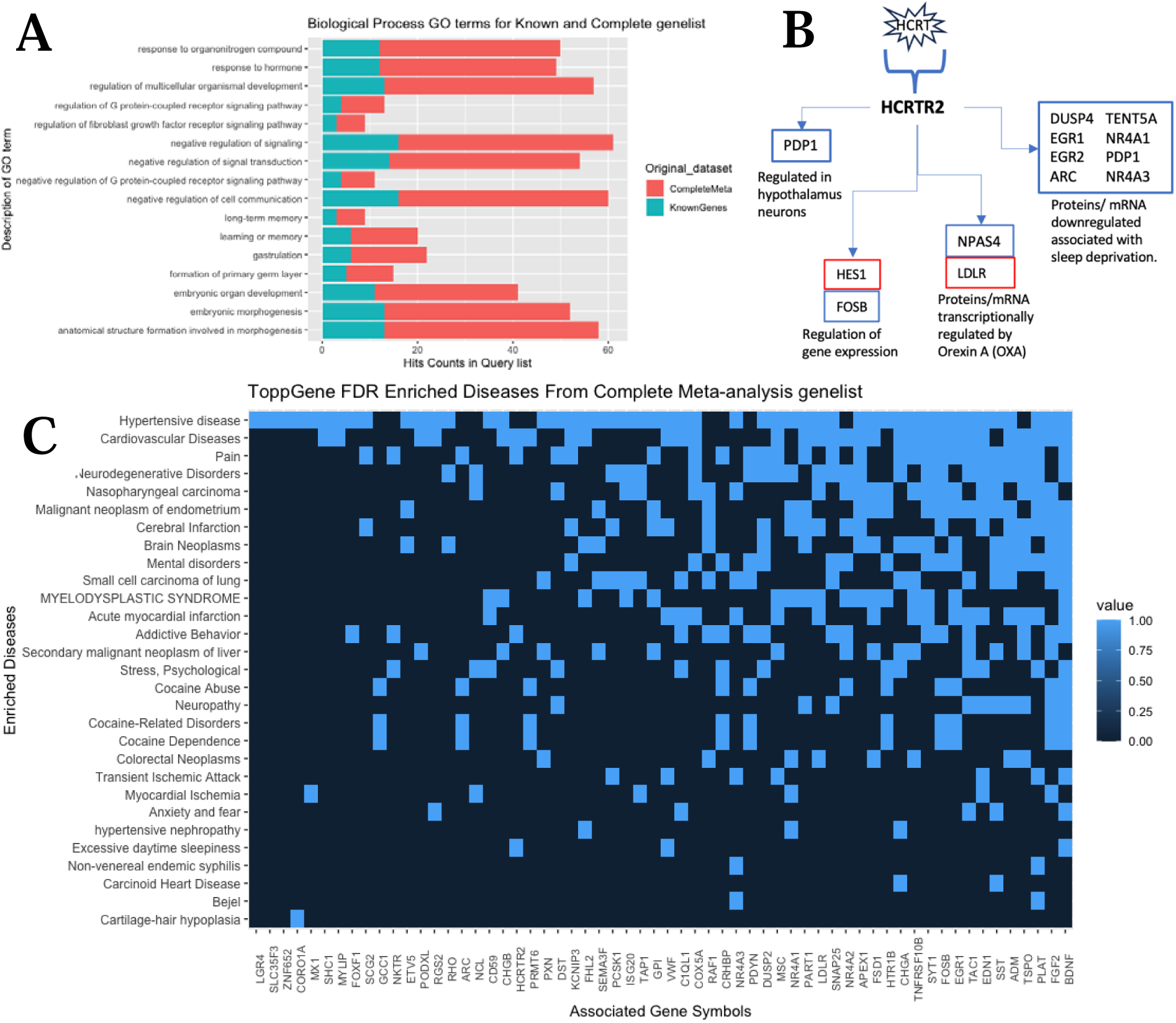
**A.** Bar plot of overlapping biological process GO terms from both complete meta-analysis gene lists (Bonferroni-adjusted p-value < 0.05) and retained genes that were previously reported in the four studies. This is addressing the 66 genes that overlap, as seen in the Venn diagram. **Figure 3B.** Only Bonferroni corrected p-value < 0.05, enriched gene pathway (orexin receptor pathway) gene list (13 genes) from complete meta-analysis gene list. Up regulation (red) and Down regulation (blue). **Figure 3C.** Heat plot of enriched disease from complete meta-analysis gene list. Presented diseases were reduced from 29 to 15 significantly enriched diseases to focus on neurological and sleep-related disorders.

#### Pathway Enrichment: Orexin

Overall functional enrichment analyses of the genes significant in meta-analysis using ToppFunn (MsigDB biocarta v7.5.1) show the orexin receptor pathway (48–50) as the only significantly enriched gene pathway (Bonferroni p-value = 8.39 x 10^-4^). The orexin receptor pathway includes genes involved in the orexin receptor signaling system that encompasses a wide range of functions such as regulating the sleep-wake cycle, energy homeostasis, neuroendocrine function, glucose metabolism, reward seeking, and drug addiction. Within the significant meta-analysis genes, two primary gene categories are present—increased expression in OOD cases of genes associated with sleep deprivation (*DUSP4, EGR1, ARC, NR4A3, NR4A1, PDP1, EGR2*) and the increased expression in OOD cases of one of two primary receptors for orexin A and orexin B (*HCRTR2*) (see conceptual model; Figure 3.B). Although these genes have a variety of functions, these categories were used in the descriptions within the framework of the orexin receptor signaling pathway. Also, it is important to note that the direction of effect seen in these genes are opposite of what is associated with wakefulness and coinsides with sleep/ sleepness phenotypes (Supplementary Figure 4).

#### Disease Enrichment

Using the meta-analysis gene list, we conducted a disease enrichment analysis to find associations between the observed DEGs and more than 11 diseases that could be linked to OOD. Using the DisGeNET database (31), we identified 29 diseases with FDR p-values < 0.05 (Figure 3.C) and 4 diseases with Bonferroni corrected p-values < 0.05: cocaine abuse (p-value = 2.29 x 10-2), stress-psychological (p-value = 4.88 x 10-2), cocaine-related disorders (p-value = 2.29 x 10-2), and hypertensive disease (p-value = 4.88 x 10-2), as seen in Table 2. When grouped based on symptom, morphology, and localization characteristics, five categories are observed: psychiatric disorders, substance abuse disorders, cardiovascular diseases, cancer diseases, and miscellaneous.

**Table 2.** Differentially expressed genes significantly enriched for a spectrum of diseases. Significantly enriched diseases that had FDR p-values < 0.05 can be seen (N = 29). Bonferroni corrected p-values < 0.05 can be seen in red (N = 4).

| Psychiatric disorders | Substance abuse disorders | Cardiovascular diseases | Cancer diseases | Miscellaneous |
| --- | --- | --- | --- | --- |
| Pain | Addictive behavior | Hypotensive disease | Nasopharyngeal carcinoma | Bejel |
| Neurodegenerative Disorders | Cocaine abuse | Cardiovascular disease | Malignant neoplasm of endometrium | Cartilage—hair hypoplasia |
| Mental disorders | Cocaine-related disorders | Cerebral infarction | Brain neoplasms | Nonvenereal endemic syphilis |
| Stress, psychological | Cocaine dependence | Myelodysplastic syndrome | Small cell carcinoma of lung |  |
| Neuropathy |  | Acute myocardial infarction | Secondary malignant neoplasm of liver |  |
| Anxiety and fear |  | Transient ischemic attack | Colorectal neoplasms |  |
| Excessive daytime sleepiness |  | Myocardial ischemia | Carcinoid heart disease |  |
|  |  | Hypertensive nephropathy |  |  |

### Genetics and Differentially Expressed Genes

#### Partitioned Heritability with Published GWAS

We examined whether DEGs associated with OOD were enriched for genetic signals associated with opioid addiction and other related phenotypes using Stratified linkage disequilibrium score regression (s-LDSC). We evaluated the 335 meta-analysis gene set against the 38 phenotype traits previously reported in Gaddis et al., 2022 (5) for opioid addiction GWAS. Given the enrichment in our DEG for the orexin pathway and its known role in sleep we added two phenotypes from Jansen et al., 2019 (36) for insomnia GWAS, three phenotypes from Dashti et al., 2019 (35) for sleep duration GWAS, and one phenotype from Lane et al., 2019 for sleep duration. After filtering for FDR adjusted p-values < 0.05, no phenotypes met the significant threshold (Supplementary Table 2).

#### Colocalization Analysis of Differential Expressed Gene and GWAS Signals

We conducted a colocalization analysis to determine whether any of the known eQTLs associated with the significantly enriched DEGs were associated with a trait or phenotype of interest. We ran the colocalization analyses using the coloc R package (42,51) on 36 phenotypic traits, some previously reported in Gaddis et al. (5) for opioid addiction GWAS and others from sleep cycle studies (Supplementary Table 5). We used eQTL single nucleotide polymorphisms from each of the 335 meta-analysis significantly enriched genes and found one significant colocalization between the DEG *FADS1* and sleep duration (posterior probability H4 = 0.0196). *FADS1* has been linked to neuropsychiatric disorders and sleep, which could be linked to the orexin receptor pathway (but not enriched in the pathway) (52).

## Discussion

The goal for this study was to extend discovery and assess the robustness of differences in gene expression associated with OOD by conducting a meta-analysis of four recently published, independent transcriptome-wide case/control studies of the DLPFC. Our meta-analysis identified 335 DEGs, of which 269 were novel and 66 were retained from the gene lists previously reported by the independent studies (6–9). Gene set enrichment analyses indicated several biological features that suggest that these DEGs dysregulate biology important to addiction. GO term enrichment results characterized OOD-associated DGE as functionally related to synaptic signaling and morphogenesis (Figure 2.A). Follow-up of DEGs in reactome annotations found enrichment in 5 signal transduction pathways (Figure 2.B). Additionally, the set of meta-DEGs were significantly enriched for the orexin pathway (Figure 3.B) and associated with several diseases across the spectrum of substance use, psychiatric disorders, cardiovascular diseases, and cancers (Figure 2.C). There was little evidence that the observed DGE resulted from differences in cell-type proportion between cases and controls, with a difference for microglia observed in one of four cohorts and differing directions of effect across cohorts. Similarly, there was limited evidence of genetic drivers associated with addiction for most of the observed DGE. Four phenotypes shared heritability with DEGs: educational attainment, schizophrenia, bipolar disorder, and Alzheimer’s disease, only in less strict significant parameters (p-value < 1.0). Colocalization and evaluation of eQTLs did not identify variants associated with DEG that were also associated with GWAS signals. These findings suggest that these observed transcriptional changes are more likely to be consequences of opioid use, rather than underlying genetic risk. Overall, our results show increasing robustness of DEG findings with increased sample size and independent cohort contribution, and they implicate the orexin pathway and other signaling within the DLPFC associated with OOD.

Figure 4 provides a conceptual model linking DEGs and enriched pathways from across our analyses to suggest a coherent hypothesis of gene dysregulation associated with OOD in DLPFC. We detail these connections below. When we investigated all DEGs using the reactome database, the five pathways that were enriched converged on *BDNF*-activated TRK receptor signaling and *FGF2* activated FGFR signaling (both nested within the RTK pathway), both of which lead to transcriptional regulation via CREB TF activation, neurite outgrowth, and plasticity (Figure 2.B & Figure 4) (53). This is important because both *BDNF* and *FGF2* has well-established associations with drug addiction (54–60). However, what is not always clear is which of the three downstream signaling cascades *BDNF* & *FGF2*-RTKs activates: (1) PLC-gamma, (2) Pi3K-AKT, or (3) MEK/ERK/MAPK signaling (53,61). Our data indicates, downstream activation of MEK/ERK signaling because *SHC1*, a DEG within the enriched pathways, binds to GRB:SOS when phosphorylated, which activates MAPK signaling (62). GPCR signaling (enriched pathway) can activate *PLC* mediated upregulation of intracellular Ca 2+, *PLC* mediated activation of MAPK via *RAS* GRP/GRP (Figure 4). TRK-MEK/ERK signaling has been shown to result in neuronal growth and plasticity (63). The observed up-regulation of *FGF2* could mediate transcriptional activation causing synaptic plasticity and growth as well (64). Together our functional enrichments of the meta-analysis DEGs show functions of RTK signaling and associated downstream events, which helps to elucidate the specific signaling cascade by which opioid overdose death, and posssibly opioid addiction, affects the neuronal plasticity within the DLPFC.

**Figure 4:**
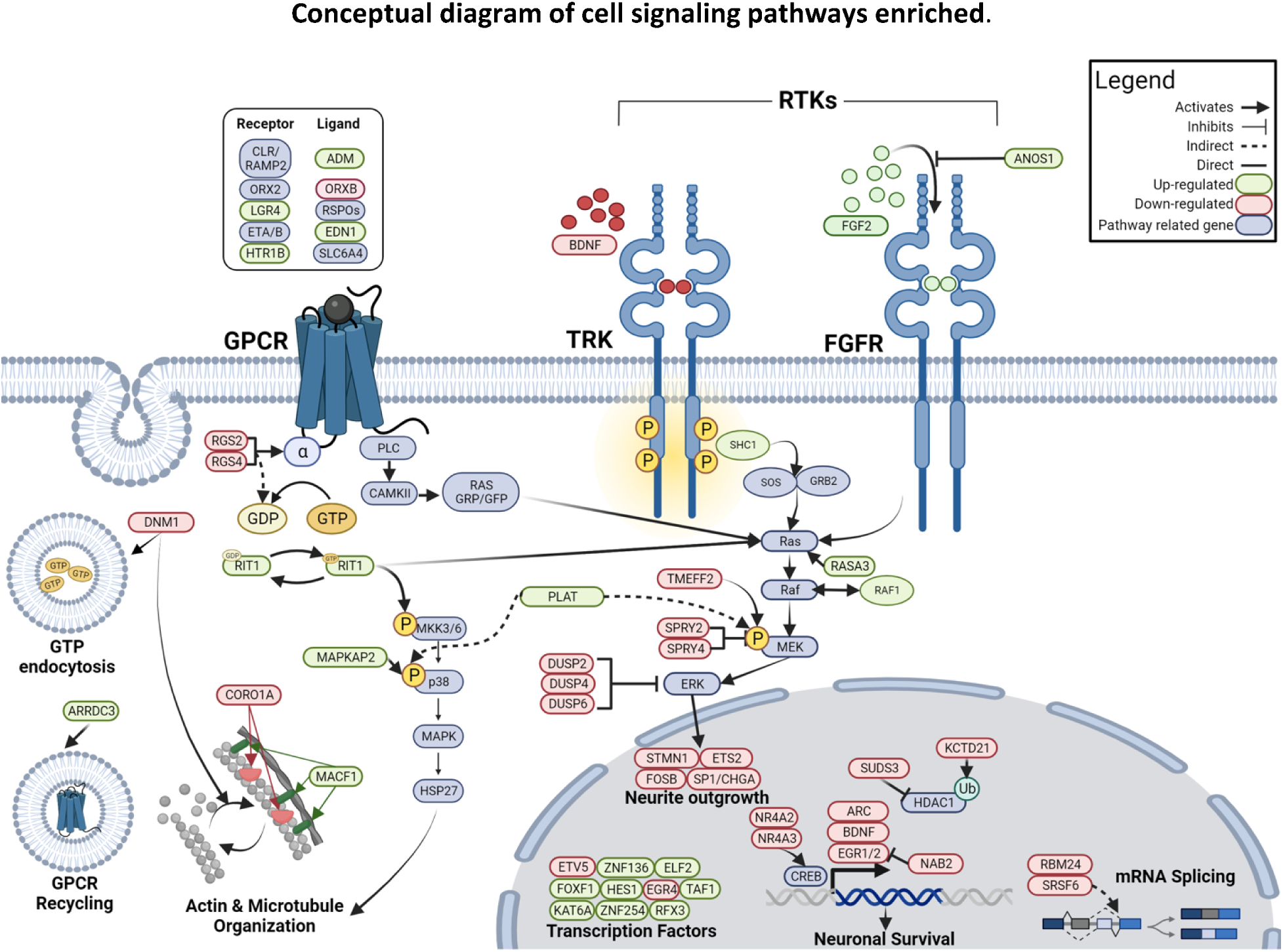
Schematic including the DEGs within the orexin receptor pathway, GPCR signaling, and Tyrosine receptor Kinase (Trk) signaling enriched using ToppFunn, Gene Ontology (GO), Reactome enrichment analyses respectively. Receptors and gene products from the hit list for each pathway are included in this depiction. Solid arrows indicate direct regulation while dashed arrows indicate indirect regulation. Genes are colored to represent significant up regulation in green and down regulation in red. Additional pathway related genes that are included are colored blue. Within this diagram we see themes of synaptic plasticity in the pathways leading to actin/microtubule organization, neurite outgrowth, and GPC receptor recycling. We also see transcriptional plasticity, which is important response in addiction through the differential expression and regulation of activity regulated TFs (BDNF, ARC, DUSP2/4/6, ETV5, EGR1/2/5), chromatin remodelers (KAT6A, SUDS3, KCTD21). Together this diagram represents potential mechanisms by which these signaling pathways may be affected by opioid overdose.

Another key finding that emerged in this meta-analysis is significant enrichment of genes in the orexin receptor pathway (MsigDB biocarta, v7.5.1). Orexin signaling occurs via activation of G-protein coupled hypocretin receptors ORX1 and ORX2. The neuropeptides orexin A (ORXA) and orexin B (ORXB) can bind and activate ORX2, whereas ORX1 can be activated by ORXA only (49). Orexin neurons are mostly located in hypothalamus, but their projections span many brain regions including the DLPFC (61,65,66). Our data show down-regulation of the ORX2 receptor, which activates MAPK and ERK signaling and regulates the sleep-wake cycle (67). Additionally, the activation of the ORX1 or ORX2 (GPCR) from ORXB (enriched DEG) mediates PLC activation which then can activate RAS GRP/GFP from the CAMKII enzyme. The *CAMK2B* gene that regulates CAMKII is nominally significant DEG in our meta-analysis (wfisher p-value = 0.00138, adj p-value = 0.0619) and is known to interact with the *ARC* and *RAS* genes (enriched DEGs), and also ERK/MAPK pathways (61,68). This activation of the *RAS* GRP/GFP can link GPCRs to *RAS* which is also a target of RTKs (Figure 4). It is important to note that orexins are dynamic with the circadian rhythm. In neurotypical brains, orexins are up-regulated during wake cycles and down-regulated during sleep cycles (67). A disruption of ORX2 has been shown in animal models to cause narcolepsy-like symptoms (69). Although orexin signaling has mainly been studied under the lens of the sleep-wake cycle, newer studies have shown associations with other neurological diseases including Parkinson’s disease (70) and addiction (71). It has been shown that patients with narcolepsy have low to undetectable orexin-producing neurons in their brains and there are low rates of substance use among narcoleptics, which is attributed to reduced activation of the mesolimbic reward circuitry (67,69). On the other hand, patients with OUD tend to have much higher levels of orexin-producing neurons, and one of the primary withdrawal symptoms of opioid use is insomnia (67). It is important to note that the decedents in this analysis are in a state of drug satiation (opioid overdose), which may explain the lower expression of genes in the orexin pathway in this study and findings of higher orexin levels in patients who are in withdrawal. Several clinical trials of Suvorexant and Lemborexant, orexin receptor agonists, will help to elucidate the relationships between orexin, opiate use, and sleep (72–75). These studies and ours suggest that orexin levels are not only correlated with sleep regulation but also the brain’s reward circuitry and specifically opioid use.

Overall, this study has shown robust evidence of RTK-mediated synaptic plasticity and significant down-regulation of the orexin signaling pathway in OOD brains. Interestingly, studies have also established associations between orexin signaling, *BDNF*, and *FGF2*. Several studies that test orexins as therapeutics for neurological diseases, including anxiety (76), depression (77), and Parkinson’s disease (70), have shown that orexin treatment is associated with the up-regulation of *BDNF*. Most notably, one study demonstrated that orexin A treatment increases *BDNF* protein in several neuronal subtypes, and orexin treatments in addition to blocking Pi3K/Akt signaling, which is known to upregulate *BDNF* transcription, did not result in BDNF up-regulation. Thus, they concluded that orexin A up-regulates *BDNF* through Pi3K/Akt signaling. This paired with our data suggests that in opioid overdose brains, orexin mediates regulation of *BDNF*, which in turn activates TRK signaling and downstream transcriptional and synaptic plasticity. GPCR signaling were also enriched with *HTR1B, EDN1, ORX2,* and ADM genes which are also associated with the orexin pathway and link to Ras and MEK/ERK/MapK signaling, from the PLC mediated activation of CAMKII (Figure 4) (61).

We employed an additional approach to characterize the DEGs by assessing their enrichment for established disease associations. This analysis revealed significant enrichment of DEGs associated with substance use disorders, specifically cocaine-related disorders, and a range of neurological disorders encompassing mental health conditions, pain syndromes, neurodegenerative diseases, and excessive daytime sleepiness. Of particular interest, a subset of enriched genes exhibited associations with multiple diseases, as illustrated in Figure 3.C Genes like *HTR1B, ARC, FOSB, FGF2, EGR1, BDNF,* and *GCC1* demonstrated overlaps in their disease associations. Unexpectedly, we also observed an enrichment of DEGs associated with cardiovascular diseases and cancer among those related to OOD. These cardiovascular conditions spanned a spectrum from brain-related disorders such as hypertensive diseases, cardiovascular disease, cerebral infarction, hypertension nephropathy, myocardial ischemia, transient ischemic attack, and acute myocardial infarction, among others.

A common thread among the majority of these diseases was disruption in blood flow and their connection to addictive responses. Given that OOD typically results from suffocation, we noted intriguing gene overlaps with diseases associated with asphyxiation. These overlaps encompassed factors such as blood flow disturbances, cerebral nutrient deficiencies, oxygen deficits in the brain, damage to the cardiovascular lining, and stroke-related processes.

### Limitations of the Study

A recurring issue within this field of OUD research using postmortem human brains is that we cannot distinguish between the effects of acute overdose death and chronic OUD: our results likely include signatures of both phenotypes. Follow-up analyses in functional paradigms with model organisms likely need to differentiate these biological processes. Although this study is the largest of DGE and OOD to date, by virtue of meta-analysis, it remains likely that it is underpowered to detect all dysregulated gene expression associated with OOD in the DLPFC. The relatively modest number of genes retained from the prior four independent cohort studies in the meta-analysis suggests the need for larger sample sizes and more diverse samples to further enhance the robustness of our gene expression findings. Another potential limitation is the use of bulk tissue RNA-seq. Single cell sequence data would potentially offer more biologically informative data within specific cell types. However, our meta-analysis of existing data has identified novel findings, despite not being sensitive to cell type–specific gene expression. Finally, the ancestral genetic heterogeneity between samples is low, consisting almost entirely of people of European descent. This does not accurately represent the population in the United States or the population of individuals with OOD. Therefore, our results miss neurobiological gene expression patterns that differ across a range of ancestral populations.

### Summary

Our primary objective was to expand our understanding of the neurobiology associated with OOD and shed light on OUD by conducting a meta-analysis of four recent, independent transcriptome-wide investigations of the DLPFC. Through this meta-analysis, we identified 335 DEGs, 269 of which were novel discoveries and 66 were identified in the prior independent studies. This meta-analysis increased the pool of DEGs associated with OOD and underscored the growing robustness of these findings with larger sample sizes and contributions from independent cohorts. Significantly, the involvement of MAPK and ERK signaling in the DLPFC through a number of pathways, including the orexin pathway, GPCR signaling, and RTK signaling, emerged as a central discovery, providing new insights into the neurobiological mechanisms of opioid overdose death and opioid addiction. DEGs exhibited significant functional enrichment in BPs related to addiction, such as synaptic signaling and morphogenesis. Furthermore, our functional enrichment analyses shed light on the signaling cascades associated with these genes, revealing RTK-mediated synaptic plasticity and a notable down-regulation of the orexin signaling pathway. These findings offer a foundation for future research into potential therapeutic interventions for OUDs by targeting the interplay between orexin (GPCR signaling), *BDNF, FGF2*, (RTK signaling) (Figure 4).

## Data Availability

All data produced in the present study are from previous published studies. All data produced in the present work are contained in the manuscript

https://www.ncbi.nlm.nih.gov/geo/query/acc.cgi?acc=GSE174409

https://www.ncbi.nlm.nih.gov/geo/query/acc.cgi?acc=GSE182321

https://www.ncbi.nlm.nih.gov/projects/gap/cgi-bin/study.cgi?study_id=phs002724.v1.p1

https://www.ncbi.nlm.nih.gov/bioproject/?term=PRJNA739548

## Acknowledgments

This work is supported by the National Institute on Drug Abuse R01 DA043980 (PIs: Scacheri, Johnson, Ackbarian), R21 DA050160 (Pls: Montalvo-Ortiz, JL), R01 DA051390 (Pls: Logan, RW), R01 DA044859 (PI Walss-Bass), and P50 DA05471 (PIs: Eric Johnson)

## Conflict of Interest

The authors declare no conflict of interest.

## Notes

### Competing Interest Statement

The authors have declared no competing interest.

### Author Declarations

Corradin O, Sallari R, Hoang AT, Kassim BS, Ben Hutta G, Cuoto L, et al. Convergence of case specific epigenetic alterations identify a confluence of genetic vulnerabilities tied to opioid overdose. Mol Psychiatry. 2022 Apr;27(4):215870. Mendez EF, Wei H, Hu R, Stertz L, Fries GR, Wu X, et al. Angiogenic Gene Networks are Dysregulated in Opioid Use Disorder: Evidence from MultiOmics and Imaging of Postmortem Human Brain. Mol Psychiatry. 2021 Dec;26(12):780312. Seney ML, Kim SM, Glausier JR, Hildebrand MA, Xue X, Zong W, et al. Transcriptional Alterations in Dorsolateral Prefrontal Cortex and Nucleus Accumbens Implicate Neuroinflammation and Synaptic Remodeling in Opioid Use Disorder. Biol Psychiatry. 2021 Oct 15;90(8):55062. Sosnowski DW, Jaffe AE, Tao R, Deep Soboslay A, Shu C, Sabunciyan S, et al. Differential expression of NPAS4 in the dorsolateral prefrontal cortex following opioid overdose. Drug Alcohol Depend Rep. 2022 Jun 1;3:100040.

