## Supplementary figures and images for "Identifying novel gene dysregulation associated with opioid overdose death: A meta-analysis of differential gene expression in human prefrontal cortex"

### Supplementary Figure 1

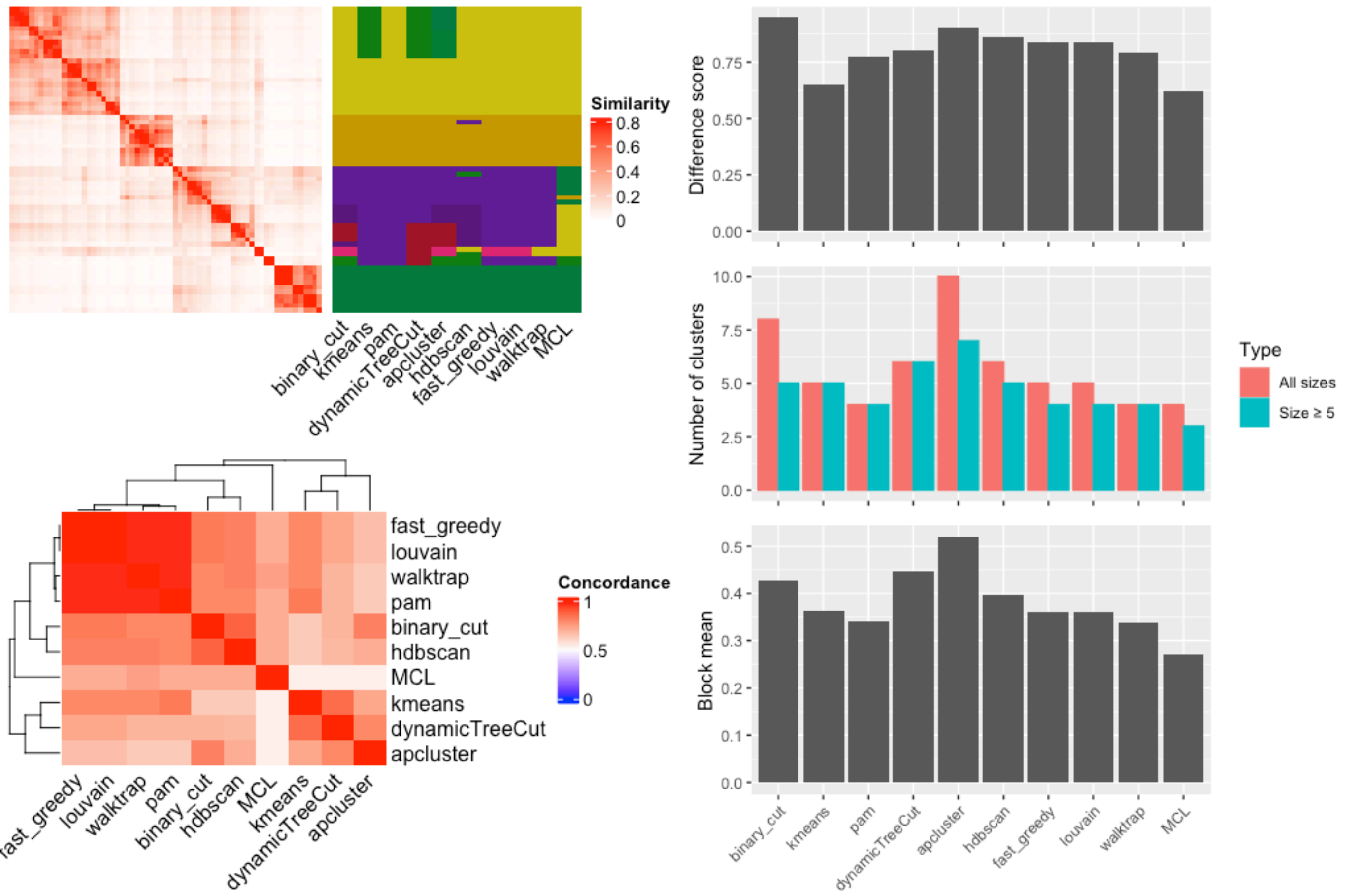
