## Supplementary Figure 2 for "Identifying novel gene dysregulation associated with opioid overdose death: A meta-analysis of differential gene expression in human prefrontal cortex"

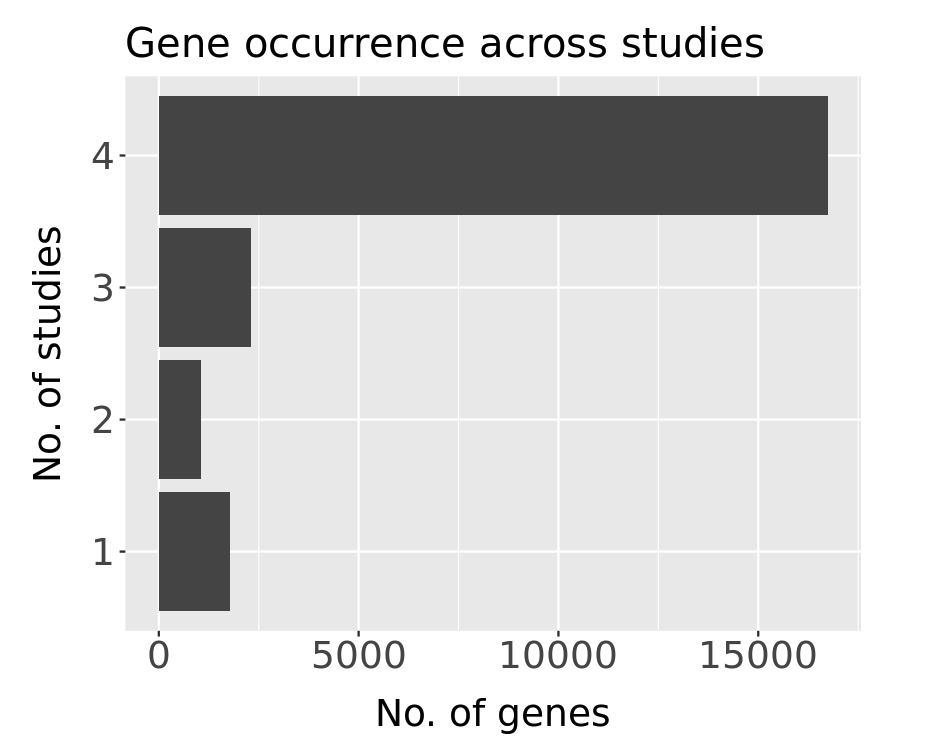

***Supplementary Figure 2.*** *Bar plot of the number of DEGs and they number of studies (of the four studies used for the meta-analysis) that they were observed in.*
