## Supplementary Figure 3 for "Identifying novel gene dysregulation associated with opioid overdose death: A meta-analysis of differential gene expression in human prefrontal cortex"

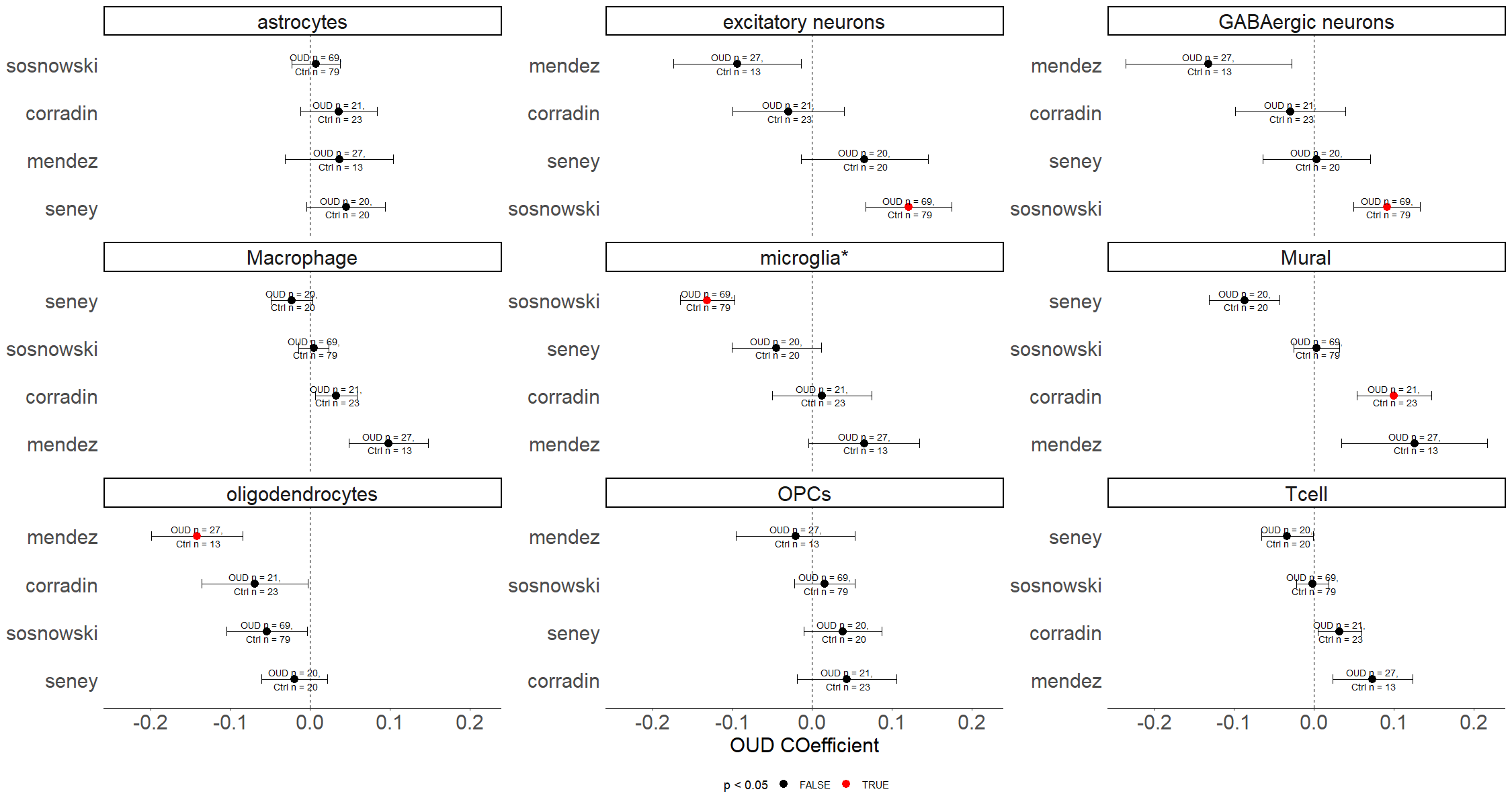

Cell type deconvolution coefficient proportions of all tested cell types on all prior studies. Red representing significant p-values <0.05

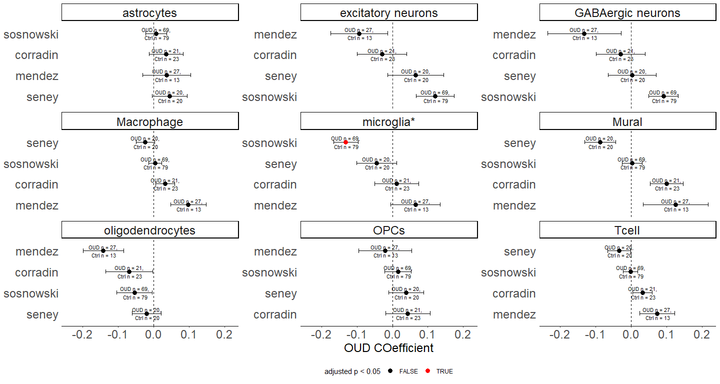

Cell type deconvolution coefficient proportions of all tested cell types on all prior studies. Red representing significant Bonferroni p-values <0.05 (FDR-BH p-values yeild same results)
