## Supplementary Figure 4 for "Identifying novel gene dysregulation associated with opioid overdose death: A meta-analysis of differential gene expression in human prefrontal cortex"

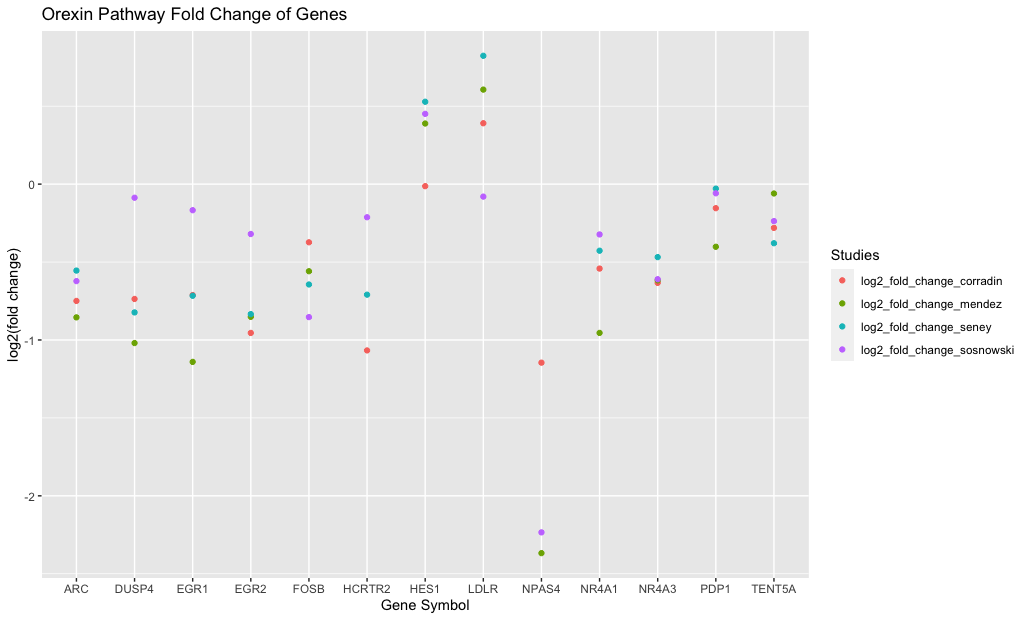

***Supplementary Figure 4:*** *Scatter plot that shows the summary stats for each case studies DGE results on the meta-analysis enriched genes for the Orexin receptor pathway.*
