## supplementary table 4 for "Identifying novel gene dysregulation associated with opioid overdose death: A meta-analysis of differential gene expression in human prefrontal cortex"

1. Alzheimer's Disease (Lambert et al., 2013 Nat Genet [24162737](https://pubmed.ncbi.nlm.nih.gov/24162737))
2. Amyotrophic Lateral Sclerosis (Rheenen et al., 2016 Nat Genet [27455348](https://pubmed.ncbi.nlm.nih.gov/27455348))
3. Autism Spectrum Disorders (Grove et al., 2019 Nat Genet [30804558](https://pubmed.ncbi.nlm.nih.gov/30804558))
4. Bipolar Disorder (Stahl et al., 2019 Nat Genet [31043756](https://pubmed.ncbi.nlm.nih.gov/31043756))
5. Cannabis Use Disorder (CUD) (Demontis et al., 2019 Nat Neurosci [31209380](https://pubmed.ncbi.nlm.nih.gov/31209380))
6. Depressive Symptoms (Okbay et al., 2016 Nat Genet [27089181](https://pubmed.ncbi.nlm.nih.gov/27089181/))
7. GSCAN Cigarettes Per Day (Saunders et al., 2022 [36477530](https://pubmed.ncbi.nlm.nih.gov/36477530/))
8. GSCAN Smoking Cessation (Saunders et al., 2022 [36477530](https://pubmed.ncbi.nlm.nih.gov/36477530/))
9. GSCAN Smoking Initiation (Saunders et al., 2022 [36477530](https://pubmed.ncbi.nlm.nih.gov/36477530/))
10. GSCAN Age of Initiation (Saunders et al., 2022 [36477530](https://pubmed.ncbi.nlm.nih.gov/36477530/))
11. GSCAN Alcohol Drinks per Week (DPW) (Saunders et al., 2022 [36477530](https://pubmed.ncbi.nlm.nih.gov/36477530/))
12. Insomnia (Jansen et al., 2019 Nat Genet [30804565](https://pubmed.ncbi.nlm.nih.gov/30804565/))
13. Insomnia (Lane et al., 2019 Nat Genet [30804566](https://pubmed.ncbi.nlm.nih.gov/30804566/))
14. Intelligence (Sniekers et al., 2017 Nat Genet [28530673](https://pubmed.ncbi.nlm.nih.gov/28530673))
15. Lifetime Cannabis Use (Ever vs. Never) (Pasman et al., 2018 Nat Neurosci [30150663](https://pubmed.ncbi.nlm.nih.gov/30150663))
16. LongSleepDur (Dashti et al., 2019 Nat Commun [30846698](https://pubmed.ncbi.nlm.nih.gov/30846698/))
17. Mean Accumbens Volume (Hibar et al., 2015 Nature [25607358](https://pubmed.ncbi.nlm.nih.gov/25607358/))
18. Mean Caudate Volume (Hibar et al., 2015 Nature [25607358](https://pubmed.ncbi.nlm.nih.gov/25607358/))
19. Mean Hippocampus Volume (Hibar et al., 2015 Nature [25607358](https://pubmed.ncbi.nlm.nih.gov/25607358/))
20. Mean Pallidum Volume (Hibar et al., 2015 Nature [25607358](https://pubmed.ncbi.nlm.nih.gov/25607358/))
21. Mean Putamen Volume (Hibar et al., 2015 Nature [25607358](https://pubmed.ncbi.nlm.nih.gov/25607358/))
22. Mean Thalamus Volume (Hibar et al., 2015 Nature [25607358](https://pubmed.ncbi.nlm.nih.gov/25607358/))
23. Total Intracranial Volume (ICV) (Hibar et al., 2015 Nature [25607358](https://pubmed.ncbi.nlm.nih.gov/25607358/))
24. Neo-conscientiousness (de Moor et al., 2012 Mol Psychiatry [21173776](https://pubmed.ncbi.nlm.nih.gov/21173776))
25. Neo-openness to Experience (de Moor et al., 2012 Mol Psychiatry [21173776](https://pubmed.ncbi.nlm.nih.gov/21173776))
26. Neuroticism (Okbay et al., 2016 Nat Genet 27089181)
27. Opioid Addiction: GENOA GWAS meta-analysis
28. Post-traumatic Stress Disorder (Nievergelt et al., 2019 Nat Commun [31594949](https://pubmed.ncbi.nlm.nih.gov/31594949))
29. Psychiatric Genetics Consortium Cross-disorder GWAS (Schizophrenia, Bipolar Disorder, MDD, ASD and ADHD) (Cross-Disorder Group of the Psychiatric Genomics Consortium, 2013 Lancet [23453885](https://pubmed.ncbi.nlm.nih.gov/23453885))
30. Schizophrenia (Ripke et al., 2014 Nature [25056061](https://pubmed.ncbi.nlm.nih.gov/25056061))
31. ShortSleepDur (Dashti et al., 2019 Nat Commun [30846698](https://pubmed.ncbi.nlm.nih.gov/30846698/))
32. sleepDuration (Dashti et al., 2019 Nat Commun [30846698](https://pubmed.ncbi.nlm.nih.gov/30846698/))
33. Sleepdur (Jansen et al., 2019 Nat Genet [30804565](https://pubmed.ncbi.nlm.nih.gov/30804565/))
34. Subjective Well Being (Okbay et al., 2016 Nat Genet [27089181](https://pubmed.ncbi.nlm.nih.gov/27089181))
35. Years of Schooling (Okbay et al., 2016 Nature [27225129](https://pubmed.ncbi.nlm.nih.gov/27225129))
